# A Three-subtype Molecular model of Cervical Cancer: Multiple PI3K Pathway inhibitors suppress growth and cooperate with HPV-directed immunotherapy

**DOI:** 10.64898/2026.01.21.26344562

**Authors:** Hong Lou, David Langan, Nick Syracuse, Elizabeth A. Murphy, Sojung Kim, Emma Robinson, Nicole M. Rossi, Yi Xie, Sonam Tulsyan, Tawnjerae Joe, Isabel Rodriguez, Nina Rao, Matthew R. Oberley, Mathias Oelke, Michael Dean

## Abstract

**Objective:** Cervical cancer is caused by human papillomavirus (HPV) infections; however, there are no molecularly defined subtypes, and few approved targeted therapies. We defined molecular subtypes and tested targeted agents.

**Methods:** Public datasets were analyzed; cell lines were treated with drugs; and donor T cells and their proliferation were measured.

**Results:** We define three molecular subtypes: I, Wild type for *PIK3CA*/no *YAP1* amplification; II, *PIK3CA* mutation/no *YAP1* amplification; III, *PIK3CA* WT/*YAP1* amplification. Patients with *YAP1*-amplified cervical cancer have poorer survival. The PI3K-specific inhibitors Alpelisib (BYL-719) and Inavolisib (GDC0077) inhibit the proliferation of multiple *PIK3CA*-mutated cervical cancer cell lines, but not a *PIK3CA* wild-type (WT) line. The pan-AKT inhibitor, Capivasertib (AZD5363), suppressed some but not all tested *PIK3CA*-mutated cell lines and one *PIK3CA*-wt cell line (SiHa). Alpelisib inhibits the expression of the HPV16 E7 oncoprotein, *CD274*/PD-L1, *YAP1*, and *EGFR* genes, only in PI3K-mutated cell lines. Treatment of an HPV16-positive, HLA-A2, *PIK3CA* mutant cell line (CaSki) with T cells (NexImmune), specific to HPV16 tumor antigens inhibited in a T cell: target cell ratio-dependent manner. BYL-719, in combination with donor T cells, enhances cytotoxicity against CaSki cells. Furthermore, pretreatment with BYL-719 and removing the drug, followed by treatment with donor T cells, had the maximum effect.

**Conclusions:** Our study revealed molecular inhibitors targeting mutant *PIK3CA* cervical cancer. When combined with immune therapies, these agents may improve outcomes of advanced HPV16 cancers. Further research on targeted therapies will improve the prognosis of patients with cervical cancer.

**Graphical Abstract:** 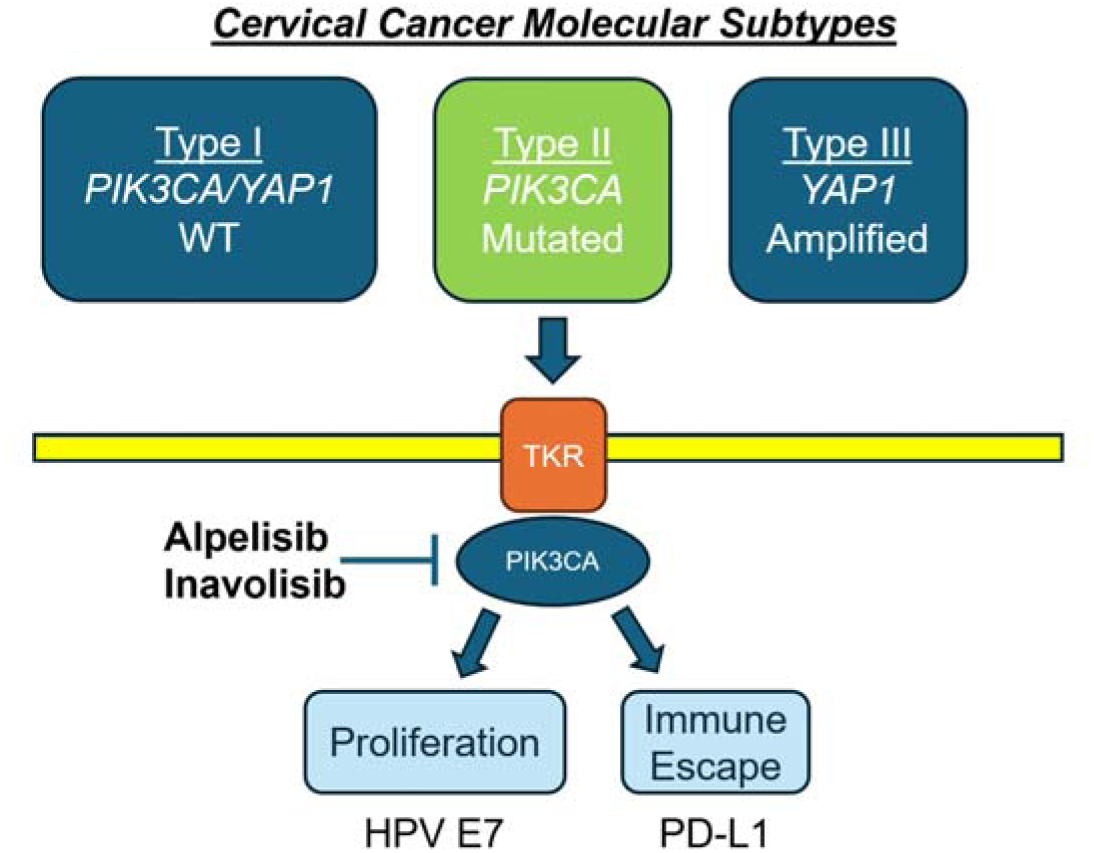

**Highlights:**

- **Identified 3 molecular subtypes of CC based on PIK3CA and YAP1 amplification status**
- **Cervical cell lines with *PIK3CA* mutation are suppressed by targeted inhibitors**
- **PI3K inhibitors Alpelisib/Inavolisib selectively block PIK3CA-mutant cells**
- **PIK3CA mutation is associated with higher expression of the checkpoint CD274/PD-L1**
- **PI3K inhibitors cooperate with donor-derived T cells to kill cervical cells**

## Introduction

Cervical cancer (CC) is the most prevalent human papillomavirus (HPV) driven tumor. While most infected individuals spontaneously clear HPV, persistent infections of high-risk (HR) HPV types in cervical tissue can lead to cervical intraepithelial neoplasia (CIN), the precursor to CC. The estimated global annual impact of CC is as high as 569,847 new cases and 311,365 deaths [1]. More than half of CC development is associated with the highly oncogenic HPV16, and the predominant histologic subtype of invasive carcinoma is squamous cell carcinoma (SCC), accounting for about 75% of cases.

HPV is a foreign pathogen expressing antigens within host cells, and the immune system frequently recognizes and eliminates infected cells. However, the virus has evolved mechanisms to evade the immune system, including upregulating CD274 (PD-L1) and CTLA-4 and silencing expression of most viral proteins in tumor cells. Immune escape is also crucial for cancer survival and metastasis [2]. Specifically, PD-L1 is overexpressed in virally induced cancers. There are high levels of PD-L1 in approximately 95% of cervical precancer and more than 80% of cervical SCC [3–5].

Clinical trials document that immunotherapy for this malignancy is feasible, and multiple new agents are currently being tested, including anti-PD1 and anti-PD-L1 antibodies [6]. Sampling of 18 tumor types showed that immune cytolytic activity corresponding to expression cytotoxic T cell genes (i.e., Granzyme A and Perforin) is higher in CC than in most other cancers, and is positively correlated with higher mutation load [7]. Furthermore, the importance of targeting neo-antigens and non-viral, tumor-associated antigens in HPV-driven cancers has been demonstrated [8].

Yes Associated Protein (*YAP1*) gene amplification and *PIK3CA* mutations are the most frequent alterations in CC [9]. Mutations in the *PIK3CA* gene are present in 13% to 46% of CC cases [10, 11]. Two specific APOBEC-induced *PIK3CA* mutations, E542K and E545K, in the helical domain of the p110 protein predominate in CC. Mutant PIK3CA proteins can support proliferation and cell survival as well as induce immune escape through PD-L1 expression. [12] [13], Alpelisib (BYL-719, Novartis Pharma AG) and Inavolisib (GDC-0077, Genentech, Inc.) are PI3Kα (PIK3CA) selective inhibitors and a clinical trial showed that *PIK3CA* mutations, are the best positive predictor of sensitivity for BYL-719 [14, 15]. BYL-719 and GDC-0077 have shown significant efficacy in human solid tumors [16], including CC [17]. The U.S. Food and Drug Administration (FDA) approved BYL-719 and GDC-0077 for the treatment of specific subtypes of breast cancer (Ref-GDC-0077) [18, 19].

AKT is a serine/threonine kinase central to the PI3K–AKT–mTOR pathway, regulating cell survival and proliferation [20]. Capivasertib (AZD-5363, Astra-Xeneca) is a potent, selective, ATP-competitive pan-AKT inhibitor (AKT1/2/3), and preclinical studies showed greater sensitivity in *PIK3CA*-mutant models [21, 22]. AZD-5363 is FDA-approved when combined with fulvestrant to treat advanced or metastatic breast cancer.

CC is highly heterogeneous in its clinical and molecular profiles [23] and lacks recognized molecular subtypes or specific targeted therapies. In this study, we classified CC into three subtypes: I). *PIK3CA* wild type, II). *PIK3CA* mutated with no *YAP1* amplification and III). *PIK3CA*-wt/*YAP*1 amplification. We investigate whether BYL-719, GDC-0077, and AZD-5363 inhibit CC cell lines with *PIK3CA* activation and whether combining BYL-719 and T cell-based immunotherapy has additive anti-tumor effect. This study provides preclinical data to support the use of BYL-719, GDC-0077, and AZD-5363 in *PIK3CA*-mutated CC.

## Results

### Establishment of three subtypes of CC

To define molecular subtypes of CC suitable for targeted therapy, we examined the distribution of the two most frequently activated oncogenes, *PIK3CA* and *YAP1* (**Figure 1A**). Analysis of 336 cervical cancers and 21 cell lines allowed for grouping of samples into three subtypes: I) tumors *PIK3CA* WT without *YAP1* amplification (2210, 65%), II) tumors with *PIK3CA* mutation without *YAP1* amplification (976, 29%), and III) tumors *PIK3CA* WT with *YAP1* amplification (180, 5.3%). Only 20 tumors and one cell line (CaSki) have both *PIK3CA* and *YAP1* activation, and *PIK3CA* and *YAP1* are significantly mutually exclusive (p<0.00001) (**Figure 1B**). A detailed distribution of the CC subtypes is shown in **Supplemental Figure 1,** and we confirmed that *YAP1* amplification is positively correlated with its expression in the Cancer Genomics Atlas (TCGA) cervical cancer (TCGA-CESC) study (**Supplemental Table 1I**) [24]. *PIK3CA* WT without *YAP1* amplification is 54-67% in five independent cohorts and 63% in 19 cervical cancer cell lines **(Supplemental Figure 1).** *PIK3CA* mutations without YAP1 amplification were observed in 27%-36% of cervical tumors and in 21% of cell lines. *YAP1* amplification frequency was 3.3-9.9% in tumors and 16% in cervical cancer cell lines (**Table 1, Supplemental Figure 1**). Including mutations in *PTEN* and *AKT1* in the Type II groups (PI3K pathway) brings the percentage of samples to 30-39% (**Supplemental Table 2**). The Caris data replicate our previous associations of *PIK3CA* mutation with older age and *YAP1* amplification, younger age at collection (**Supplemental Table 3**), and a higher frequency of *YAP1* amplification in squamous cell carcinoma (χ2 = 18, P < 0.00001). TCGA tumors with *YAP1* amplification (subtype III) adjusted by clinical stage were significantly associated with poorer survival outcomes in TCGA-CESC (HR=3.2 (95% CI 1.5, 8.0) (**Figure 1C**). We replicated this association in Caris, and after adjusting for age and race, *YAP1*-amplified subjects have poorer overall survival compared to patients without *YAP1*-amplified tumors (HR=1.4 (95% CI 1.1-1.8), (**Figure 1D**).

**Figure 1.**
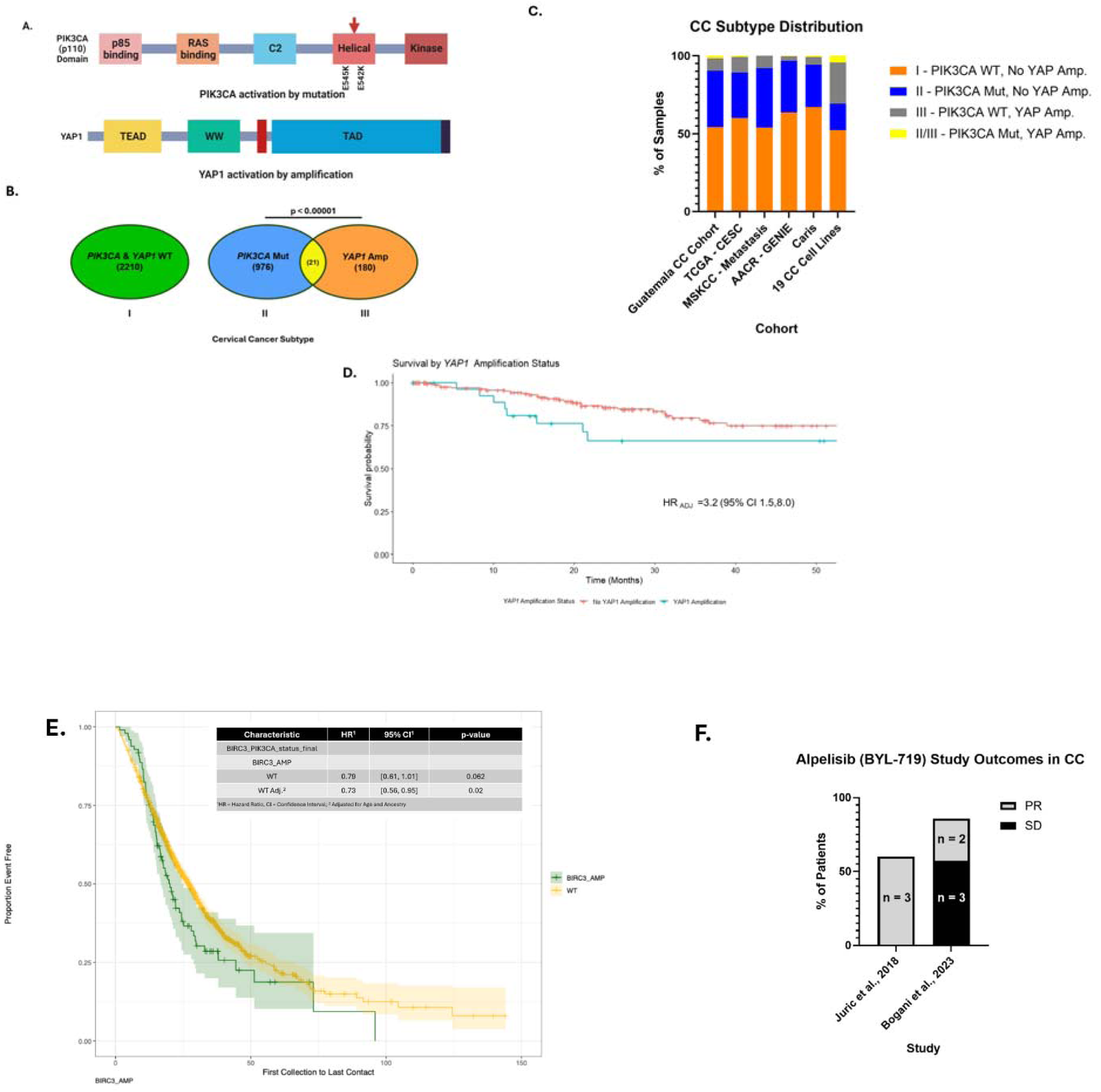
Modular structures of the two most prevalent oncoproteins, p110 (*PIK3CA)* and YAP1 in CC development. (A) Somatic mutations in the helical domain of *PIK3CA,* activating the p110 catalytic subunit of PI3K, and *YAP1* amplification occur frequently in CC. (B) *PIK3CA* mutations and *YAP1* amplification are significantly mutually exclusive. The numbers represent the combined data from Guatemala, TCGA-CESC, MSKCC, and AACR-GENIE datasets. (C) Distribution of three molecular subtypes of cervical cancers. (Complete data is in Supplemental Table 1.) (D) TCGA CC patients with *YAP1* amplification, adjusted for clinical stage, had significantly worse survival outcomes than those without *YAP1* amplification. (E) Survival curve for *YAP1*-amplified CC in the Caris dataset. (F) Cervical cancer patient outcomes from completed Alpelisib (BYL-719) trials. The respective study and year are listed. PR = Partial Response; SD = Stable Disease.

Of the three subtypes we have defined, there is an FDA-approved targeted therapy for only type II, *PIK3CA*-mutated tumors. In previously published clinical trials of BYL-719/Alpelisib, small numbers of *PIK3CA*-mutated cervical cancer patients were included. We identified two such studies, treating a total of 17 patients, and 5/17 patients displayed a partial response, and the remaining 12/17 patients had stable disease [17, 25]. Therefore, human *in vivo* data support a role for PI3K inhibitors in this subtype of CC.

### BYL-719 inhibits proliferation in CC cell lines harboring *PIK3CA* mutations

We hypothesized that specific inhibitors of mutant *PIK3CA* might suppress proliferation in CC cell lines harboring PIK3CA mutations. We tested the PI3Kα (*PIK3CA*/p110) specific inhibitor, BYL-719, in CC cell lines with and without *PIK3CA* mutations and including HPV-independent, HPV16 and HPV68-driven cells)(**Table 2**). Cells were treated with BYL-719 in an impedance-based proliferation assay, and the growth at 24h and 48h was determined. Initial studies in CaSki and SiHa cells were performed with 0-50 μM drug, and subsequent experiments with 0-10 or 0-20 μM (**Supplemental Table 3**). BYL-719 significantly inhibited cell proliferation 55-95% at concentrations as low as 5µM in all *PIK3CA*-mutated cells tested but had no significant effect on SiHa cells that are WT for *PIK3CA (***Figure 2A, Supplemental Figure 2**). From the curves, IC values were calculated, ranging from 1.3 to 15 μM (**Table 2**). For comparison, the unbound C_max_ plasma BYL-719 in patients treated with the standard dose of 300mg QD was 0.7 μM (https://www.accessdata.fda.gov/drugsatfda_docs/label/2024/212526s009lbl.pdf).

**Figure 2.**
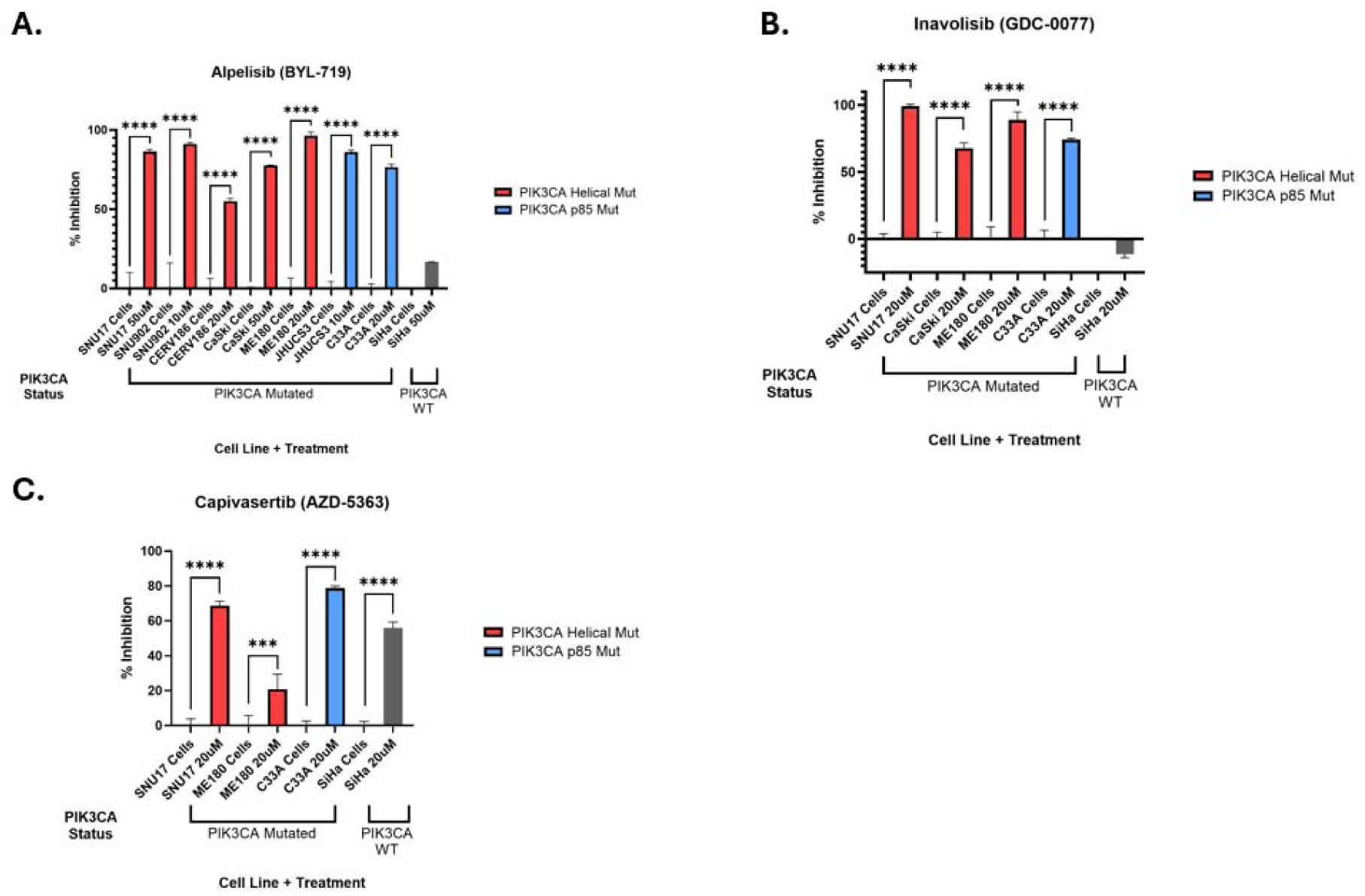
Inhibition of cervical cancer cell lines by PI3K pathway inhibitors. The maximum dose for each cell line was selected, and the percentage inhibition was compared with the corresponding control. (A) Alpelisib (BYL-719). (B) Inavolisib (GDC-0077). (C) Capivasertib (AZD-5363). All cell lines have *PIK3CA* mutations except SiHa. **** = p < 0.0001 by one-way ANOVA and Dunnett’s multiple comparison test. <20% inhibition interpreted as no or negative response.

**Table 2.** Response of cell lines to PI3K and AKT inhibitors. The half maximal inhibitory concentration (IC□□IC□□) was calculated for each cell line drug combination. Cell lines treated with BYL-719 (Alpelisib), GDC-0077 (Inavolisib), and/or AZD-5363 (Capivasertib) and assessed for proliferation. IC□□IC□□ values were calculated at fixed time points of maximal growth for each cell line. CI = Confidence Interval. For results with an IC□□IC□□ < 5 μM (the lowest tested dose), IC□□IC□□ confidence intervals are not estimable because the 50% inhibitory response was not achieved within the tested concentration range. *CI truncated at 0; delta-method lower bound < 0. * *For results with an IC50 <5uM (lowest tested dose), IC50 confidence intervals are not estimable because the 50% inhibitory response was not reached within the tested concentration range +HeLa data is from [26, 27].^MS751 data is from [27].

| Cell Line | HPV Type | PIK3CA Status | Domain | Alpelisib (BYL-719) IC<sub>50</sub> | Inavolisib (GDC-0077) IC<sub>50</sub> | Capivasertib (AZD-5363) IC<sub>50</sub> | Reference(s) |
| --- | --- | --- | --- | --- | --- | --- | --- |
| CaSki | HPV16 | E545K | Helical | 15 uM | 7.3 uM | No response |  |
| SNU-17 | HPV16 | E545K/E726K | Helical | 1.3 uM | 0.9 uM | 3.7 uM |  |
| C33A | None | R88Q | p85 | 10 uM | 8.9 uM | 1.6 uM |  |
| CERV186 | HPV16 | E545K | Helical | 2 uM |  |  |  |
| SNU902 | HPV16 | Q546L | Helical | 2.1 uM | 15.2uM |  |  |
| ME180 | HPV68 | E545K | Helical | <5 uM* | <5 uM* | 1.8uM |  |
| JHUCS-3 | None | R88Q | p85 | 1.4 uM |  |  |  |
| SiHa | HPV16 | WT |  | No response | No response | 9.2 uM |  |
| HeLa | HPV18 | WT |  | No response<sup>+</sup> |  |  | [25] |
| MS751 | HPV45 | WT |  | No response<sup>^</sup> |  |  |  |

To test an additional specific PI3K inhibitor, we treated selected cell lines with 0-20 μM of Inavolisib (GDC-0077). Three *PIK3CA*-mutated cells were strongly inhibited at all tested doses, with IC values of <5-8.9 uM (**Figure 2B**, **Table 2, Supplemental Figure 3**). As with BYL-719, GDC did not inhibit SiHa cells, which are WT for *PIK3CA*, but instead elicited a small but significant stimulation (**Figure 2A, Supplementary Figure 3**). Finally, to determine if inhibition downstream of PI3K has an impact, we treated selected cell lines with 0-20 μM Capivasertib (AZD-5363), a pan AKT inhibitor. Two *PIK3CA*-mutated cell lines, C33A (R88Q) and SNU-17 (E545K/E726K), were strongly inhibited by AZD-5363 with IC of 1.5-3.7 μM (**Figure 2C**, **Table 2, Supplemental Figure 4**). ME180 cell (E545K) displayed a small but significant stimulation over the initial 20 h and ceased proliferation compared to the control (**Figure 2C, Supp fig2).** Surprisingly, SiHa cells (*PIK3CA* WT) were strongly inhibited by AZD-5363 with an IC of9.2 μM (**Figure 2C**, **Table 2, Supplemental Figure 4**). Therefore, we have identified multiple PI3K inhibitors that suppress the proliferation of *PIK3CA*-mutant cervical cell lines.

### Alteration of the expression of the immune checkpoint molecule *CD274* in CC and CaSki cells

To improve the efficacy of therapies for CC subtypes, a better understanding of the relationship between immune evasion and *PIK3CA* somatic mutations is needed. We analyzed the relationship between *CD274* (PD-L1) and *PIK3CA* gene expression. *CD274* expression levels in patients with a *PIK3CA* mutation (MUT) and *PTEN* wild type (WT) (n=55) are significantly higher than those of *PIK3CA* WT tumors (n=167)(P=0.008) (**Supplemental Figure 5A)**. To test the effect of *PIK3CA* mutation on PD-L1 expression, we selected CC cell lines with or without *PIK3CA* mutation (**Table 1**) and measured PD-L1(*CD274*) mRNA and protein expression. We found that PD-L1 expression in HPV16 CaSki harboring the E545K mutation is highly elevated compared to cell lines that are either WT (HPV16 SiHa) or E545K in cell lines with non-HPV16 types (HPV68 ME180) (**Supplemental Figure 5B, C).** This result suggests that the *PIK3CA* E545K mutation, combined with HPV16 in SCC tumors, contributes to immune evasion.

### BYL-719 inhibits PD-L1, YAP1, and YAP1 targets in a *PIK3CA* mutated CC cell line

To examine the impact of BYL-719 treatment on protein expression, we treated cells with 5 and 10 μM BYL-719 for 24 and 48 hours. Treatment with the PI3Kα (*PIK3CA*/p110) inhibitor, BYL-719 resulted in decreased expression of PD-L1, YAP1, EGFR, CTGF, Integrin, and HPV16 E7 protein levels in CaSki, ME180, and SNU-17 cells (**Figure 3, Supplemental Figure 6**). For CaSki and SNU-17 cells, there was an 80% reduction of the HPV E7 protein. ME180 cells harbor HPV68, and we lacked an antibody against the HPV68 E7 protein. However, BYL-719 treatment resulted in reduced levels of p16, a marker for HPV E7 expression^18^ (**Figure 3, Supplemental Figure 6B**). **Figure 3F,G** shows typical proliferation plots of a responsive and non-responsive cell line. Therefore, BYL-719 inhibition of cervical tumor cells is active at 5uM and functions in part by reducing the expression of the HPV E7 oncoprotein, and the inhibition of PD-L1 may potentiate immune therapies.

**Figure 3.**
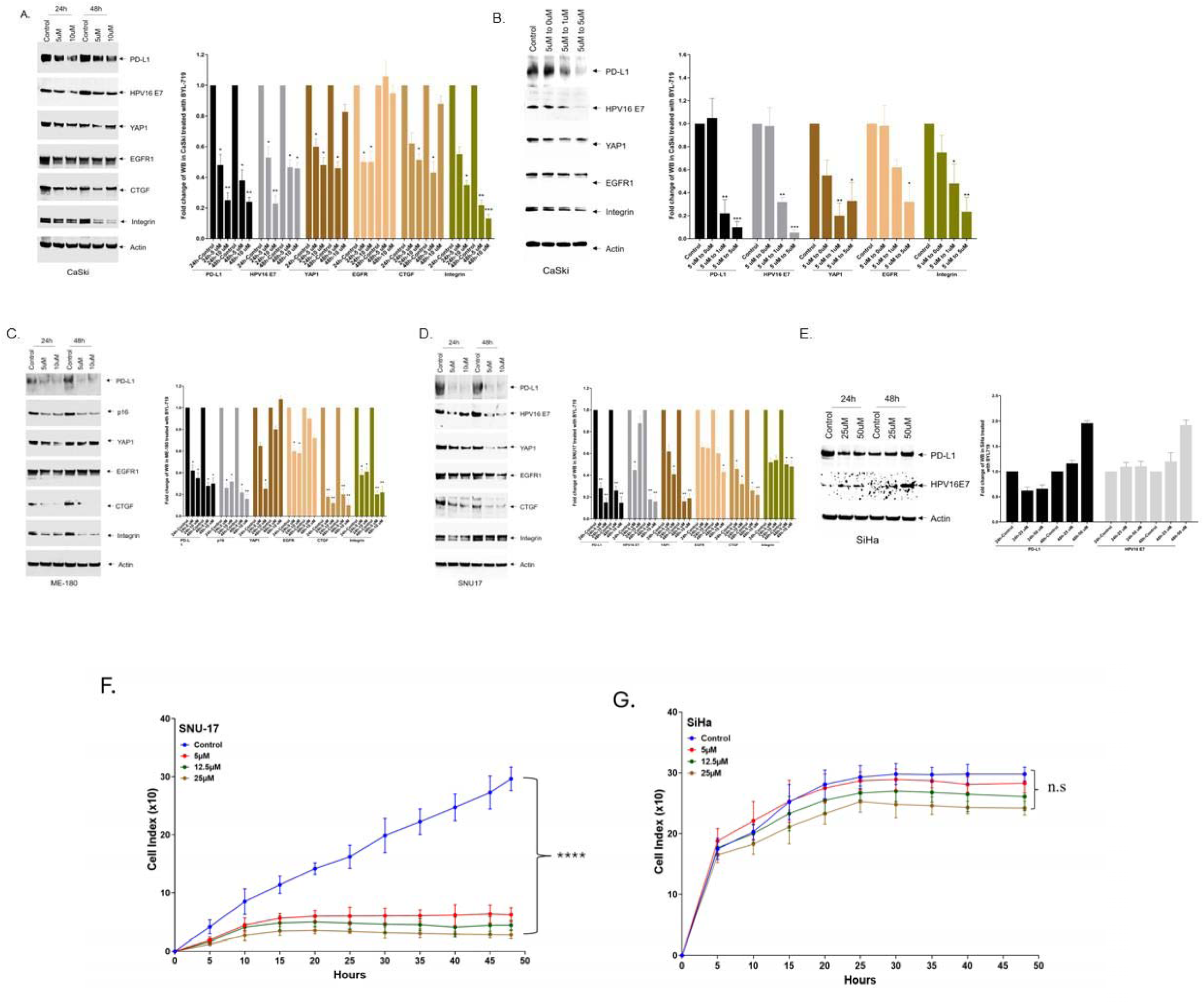
Impact of BYL-719 on the expression of target proteins in cervical cancer cell lines. (A, B) CaSki cells treated with BYL-719 show reduced levels of PD-L1, YAP1, EGFR, CTGF, Integrin, and HPV16 E7. In Panel B cells were treated with 5 uM BYL-719, washed at 24h and place in 0, 1 or 5 uM drug. (exposure time, 2 min). (C) BYL-719-treated ME180 cells show decreased levels of PD-L1 and CTGF. (D) SNU-17 cells also show reduced levels of these proteins after treatment (exposure time, 5 min). (E) SiHa (PIK3CA wild-type) shows no reduction in PD-L1 or HPV16 E7 after treatment with BYL-719 (exposure time, 3 minutes). (F) Cell proliferation plot of SNU-17 cells treated with BYL-719 (G). SiHa cells treated with BYL719. *=P<0.05, **=P< 0.01, ***=P<0.001.

### BYL-719 inhibits HPV16 and host immune genes in CaSki cells

To determine the impact of BYL-719 treatment on responsive cell lines, we performed transcriptome analysis of CaSki cells treated with BYL-719. BYL-719 inhibition resulted in a 50% -70% reduction in expression of the HPV16 E6 and E7 genes at 24h and 48h (**Supplemental Figure 7A**). Cellular immune system-related genes involved in antigen presentation (*B2M)*, or antigen processing (*TAP1*) were also downregulated (**Supplemental Figure 7B**). In addition, genes involved in the upregulation of *CD274* (PD-L1), such as *STAT1*, *IRF1[28]*, *ENO1[29]* and *NFkB[30]* were also inhibited (**Supplemental Figure 7B**). BYL-719 also inhibits the expression of the anti-viral *APOBEC3* family genes (**Supplemental Figure 7B**). In addition, BYL-719 inhibits the *YAP1*-related HPV regulatory genes[31], including *EDN1*[31], *TEAD2*[32], *NEGR1*, *TGFB1*, and *SNAI*[33] (**Fig. S7B**), which are associated with cancer cell proliferation. To validate the transcriptome analysis, we performed qPCR on selected genes, yielding results similar to those obtained from the transcriptome analysis (**Supplemental Figure 7B**). We conclude that inhibition of the PI3K pathway in *PIK3CA-*mutated CC impacts the expression of the HPV oncogenes and several human genes involved in the immune response.

### PI3Kα (*PIK3CA*/p110), BYL-719 inhibition cooperates with activated T cells to kill CC cells

To test whether BYL-719 inhibition cooperates with HPV-tumor antigen-specific CD8+ T cells, we treated CaSki (HPV16, *PIK3CA* E545K, HLA-A*02:01 SCC) cells with the drug and activated T cells, alone and in combination. Antigen-specific CD8+ T cells restricted to the common human leukocyte antigen (HLA-A*02:01) serotype were generated at research scale using the AIM ACT platform as a potential treatment for HPV-driven diseases (Nexlmmune, NEXI-003) [32]. BYL-719 was combined with CD8+ T cells generated from healthy donors, and the effect was measured using a tumor cytolysis assay. Combining BYL-719 with antigen-specific CD8+ T cells inhibited CaSki cell growth to a greater extent than either treatment alone with three of the four batches of T cells (**Figure 4 and Supplemental Figure 8A, B**) In one donor, T cells alone caused nearly maximal tumor cell killing, with the combination of these T cells and BYL-719 not significantly different than the effect of T cells alone. BYL-719 alone inhibited the T cell response in a dose-dependent manner, as measured by intracellular cytokine staining of T cells activated with TransAct beads and cultured with or without BYL-719. Even so, the combination treatment led to greater cytotoxicity of CaSki cells than either single treatment alone (**Supplemental Figure 8C**). To avoid potential T-cell inhibition by BYL-719 during cytolysis, we performed a washout experiment. CaSki cells were treated with 5µM BYL-719 for 24h to downregulate tumor cell defenses, followed by a refreshing of medium with T cells (E:T = 4:1) in no drug or 1µM BYL-719. T cells showed rapid killing of CaSki cells pretreated with BYL-719, which was significantly greater than that observed with BYL-719 pretreatment alone and comparable to the effect on untreated cells (**Figure 4C**). Maintaining a low dose of BYL-719 (1uM) did not impair T cell toxicity, demonstrating that pre-conditioning of tumor cells with PI3Ka inhibition followed by T cell therapy yielded optimal cytotoxic effects.

**Figure 4.**
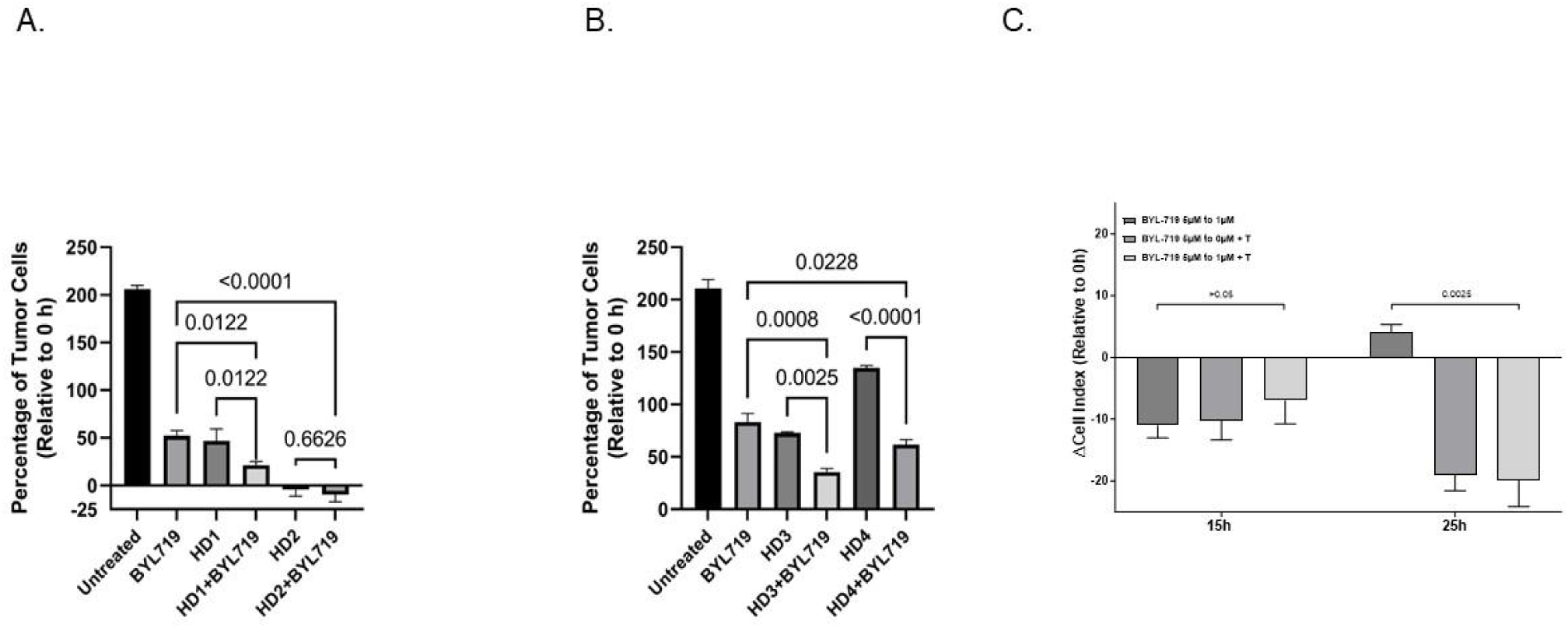
The effect of BYL-719 and CD8+ T cells on tumor cell growth. CaSki cells were cultured alone, with either BYL-719 (5µM) (A and B) or BYL-719 (1µM) (C) with T cells (E:T ratio – 2.5:1-10:1), or with BYL-719 plus T cells for 48h or 24h, respectively. We performed three independent experiments testing four batches of T cells, HD1 and HD2 in (A); and HD3 and HD4 in (B). In Figure 5C, a representative combining killing assay with the T cells (L437 HP) plus BYL-719 shows T cells combining 1µM of BYL-719 synergistic inhibitory effect compared to either BYL-719 or T cells alone. The differences (Mean ± SEM) in the percentage of CaSki cells proliferating are relative to 0 h. Each condition was run in replicates for all experiments. The rate of CaSki tumor cells relative to 0h (time of drug addition) is reported. Statistical difference was tested by a one-way ANOVA with Holm-Šidák post-test and the p-values shown.

To summarize our findings in the context of previous data, we present a model of CC evolution and potential targeted therapeutic strategies (**Figure 5**). In cervical epithelial cells infected with HPV, APOBEC3 and HPV E7 are upregulated, which can increase specific host gene mutations, e.g., *PIK3CA* E545K [33]. These genetic alterations enhance PD-L1 expression, contribute to drug resistance, and facilitate immune evasion. Our study indicates that a PI3K-specific inhibitor can effectively suppress CC cell proliferation by targeting HPV E7 and reducing PD-L1 levels. Moreover, our observations of BYL-719 alone or in combination with CD8+ T cells suggest enhanced efficacy in eradicating CC cells.

## Discussion

Prior research has shown that CC exhibits significant clinical and molecular heterogeneity [23]. However, unlike many other cancers, there are no established molecular subtypes for targeted therapy. *YAP1* amplification and *PIK3CA* mutation are the most common oncogene activation events in CC [10, 11]. We show that two genetic alterations are significantly mutually exclusive (p<0.00001), suggesting that they may drive parallel oncogenic pathways or confer similar advantages to CC development. Research on *YAP1* amplification and *PIK3CA* mutations in CC is limited. However, their interconnected roles in HPV16 cervical cancers highlight the complexity of cancer development and the potential for targeted therapies addressing these specific genetic and signaling pathway aberrations. In the current study, we categorized 1407 CC samples from Guatemala, TCGA, Memorial Sloan-Kettering Cancer Center) MSKCC and the American Association of Cancer Research (AACR_GENIE) studies, and further validated with 2284 samples from Caris. We delineated three subtypes: Subtype I: *PIK3CA* wildtype with no *YAP1* amplification; Subtype II: *PIK3CA* mutation with no *YAP1* amplification; and Subtype III: *PIK3CA* WT and *YAP1* amplification. However, all HPV-positive tumors are driven by the E6 and E7 oncoproteins, which act in all three subtypes.

We chose the *PIK3CA* mutated Subtype II to initiate preclinical validation and Alpelisib (BYL-719), Inavolisib (GDC-0077), and Capivasertib (AZD-5363) as target drugs, as these agents have shown efficacy against *PIK3CA-*mutated cancers and are FDA-approved for use in selected breast cancers with *PIK3CA, PTEN,* or *AKT1* mutations [14]. We demonstrated that the PI3K inhibitors BYL-719 and GDC-0077 selectively inhibit the growth of Subtype II (*PIK3CA*-mutated) cervical cell lines *in vitro,* with minimal effects on a *PIK3CA* WT cell line, consistent with PI3K pathway dependency. We tested five independent cell lines with mutations in the PIK3CA helical domain, the region of the gene most frequently altered in cervical cancer. In these mutant cell lines, we show that PIK3CA inhibition also reduced the expression of the HPV E7 oncoprotein and immune checkpoint protein CD274/PDL1, in addition to other oncogenic proteins such as YAP1 and EGFR.

Furthermore, the combination of BYL-719 and HPV-targeted T cells enhanced tumor cell killing *in vitro;* however, care must be taken to avoid dampening T-cell activity, as treatment with BYL-719 alone had an inhibitory effect on T-cell activity. Recently, Jiang et al studied the effect of BYL-719 on CaSki and SiHa cells and obtained IC values comparable to ours. In cell-derived xenografts (CDX), they show that BYL-719 inhibited CaSki CDXs and cooperated with an anti-PD-1 antibody. [27] A 2018 study of BYL-719, conducted in a first-in-human clinical trial involving 134 cancer patients, reported partial responses in 7 patients [17]. These included three out of five patients with cervical cancer and four with other cancers. Specifically, cervical cancers accounted for 43% of partial responses, whereas cancers of each type accounted for 14%. Additionally, the disease control rate was highest in cervical cancer patients, achieving 100% (five out of five). In addition, Bogani et al. treated 17 consecutive patients with CC with BYL-719. Six of these patients had *PIK3CA* mutations, and all had either a partial response or stable disease [25].

Our results are consistent with prior reports, showing that *PIK3CA* WT cell lines (SiHa and HeLa) do not respond to BYL-719 (Alpelisib), whereas introducing *PIK3CA* hotspot mutations (E542K or E545K) into SiHa cells via plasmid transfection confer responsiveness and sensitivity to Alpelisib (BYL-719) [26, 34]. In our study, the PI3Kα inhibitor Inavolisib (GDC-0077) showed a similar mutation-dependent activity profile. Inavolisib selectively targets PIK3CA, which encodes the PI3Kα catalytic subunit p110α where pathogenic hotspot mutations cluster in the helical (exon 9; E542K, E545K) and kinase (exon 20; H1047R) domains. Notably, Alpelisib (BYL-719, PIQRAY) has been FDA-approved since 2019 for HR+/HER2–, *PIK3CA*-mutated advanced or metastatic breast cancer in combination with fulvestrant, and in 2024, Inavolisib (GDC-0077, Itovebi) was approved in combination with palbociclib and fulvestrant for adults with endocrine-resistant, HR+/HER2–, *PIK3CA*-mutated, locally advanced or metastatic disease. Moreover, the finding that Alpelisib (BYL-719) treatment leads to reduced expression of *CD274*/PD-L1 suggests that this drug may potentiate the recognition and killing of CC (or other tumors) cells by activated T cells. Indeed, amplification of a region including PD-L1 (*CD274*), is positively associated with cytolytic activity in cervical cancer, so reducing its expression may potentiate tumor cell killing by an already established anti-tumor T cell response [7].

. Capivasertib (AZD-5363, AstraZeneca Pharmaceuticals) is a selective, ATP-competitive inhibitor of AKT that blocks its catalytic activity, thereby reducing phosphorylation of AKT substrates (e.g., PRAS40, GSK3β) and downstream mTORC1 effectors (S6, 4E-BP1). [35]. Capivasertib (AZD-5363, Truqab) is FDA-approved in combination with fulvestrant for HR+/HER2− advanced breast cancer harboring *PIK3CA*, *AKT1*, or *PTEN* alterations. The pan-AKT inhibitor AZD-5363 inhibited three out of five PIK3CA mutated cervical cancer cell lines and the only tested *PIK3CA* WT line. It has been shown that PIK3CA helical domain mutations can signal through AKT-independent pathways (Supplemental Figure XX A,{Lou, 2015 #1235}{Zhao, 2010 #1169} The use of Capivasertib in cervical cancer remains investigational, and the drug has been evaluated in early phase I solid-tumor cohorts that included cervical cancer (NCT01226316).

To address the potential for combination of PI3K inhibitors with immunotherapy we treated a cervical cell line (CaSki) with BYL-719 alone or in combination with T cells specifically activated to recognize HPV E6 and E7 peptides, as well as a tumor-associated antigen BIRC5 peptide, restricted by HLA A*02:01. Using T cells from four different donors, we were able to show that the combination of BYL-719 and activated T cells led to the most effective killing of tumor cells. We did not find that the combination was more than additive and demonstrated that BYL-719 may reduce the activity of activated T cells. However, pre-treating cells with BYL-719 and adding activated T cells in no or a low dose of the drug produced high cytotoxicity. Therefore, there is potential for using BYL-719 in combination with T-cell therapy.

Pembrolizumab (Keytruda) is approved for use in combination with chemotherapy to treat advanced CC patients who express the PD-L1 protein based on data from a large phase 3 trial (KEYNOTE-826). A phase 1 clinical trial of T cells engineered with a T cell receptor targeting HPV16 E7 (E7 TCR-T cells) for the treatment of refractory metastatic HPV-associated epithelial cancers, including patients with resistance to anti-PD-1 inhibitors, resulted in robust tumor regression [36]. Given the widespread use of anti PD-L1 therapies, it would be worth exploring PI3Ka inhibitors to potentially combat resistance mechanisms. Our combination immunotherapy experiments also suggest that the timing and mechanism of these therapies are important to optimize T cell function and effectiveness.

*YAP1* amplification characterizes subtype III of CC, and we have recently shown that this represents an aggressive subtype of CC with an early onset that is more common in patients of African descent [36]. Here, we show that patients with *YAP1*-amplified CC have poorer survival with higher mortality, even in stage 1 patients. This is consistent with *YAP1* amplification leading to enhanced tumor aggressiveness and stemness [37]. YAP1 activates super-enhancers of the *EGFR* gene, a downstream target of *YAP1*, further contributes to the proliferation, migration, and invasion of cancer cells in CC [38]. Targeted therapy for this subtype would be highly beneficial, and specific YAP1 inhibitors are currently in development. [39]. We suggest that the inhibition of EGFR alone or by antibody drug conjugates (ADC) be explored in YAP1-amplified CC (Supplemental Figure xxB). We have characterized a panel of CC cell lines and identified five with *YAP1* amplification that can be used as models for preclinical development [37].

Subtype I cervical tumors are principally driven by HPV E7. However, E7 localizes to intracellular compartments and therefore cannot be targeted by antibodies or chimeric antigen receptor (CAR) T cells. Hence, immunotherapy approaches targeting the E7 protein have promise for this subtype of CC [32, 36].

Our study has several limitations, including the small number of cell lines (only one *PIK3CA* WT cell line) and the need for *in vivo* studies. In this study, we did not address the mechanism by which PI3K regulates HPV E7 or CD274/PD-L1. However, BYL-719, GDC-0077, AZD-5363, and multiple immunotherapy approaches are already proven safe and effective. It is reasonable to propose a clinical trial. of PI3K inhibitors in *PIK3CA-mutated* CC or other gynecological tumor types alone or combined with immunotherapy. We could only test one cell lines with donor T cells, as from all available cervical cancer cell lines, only CaSki, has a *PIK3CA* mutation, HPV16, and an HLA-A02 allele to allow treatment with patient T cells.

In summary, we propose an initial molecular classification of cervical tumors to allow the development of targeted therapies and provide a plausible therapeutic strategy for *PIK3CA-mutated* (Subtype II) cervical tumors, which represent about 30% of all invasive cervical cancers. The separation of CC patients by molecular alterations could lead to improved therapeutic outcomes.

## Methods

### Cell culture and Alpelisib/BYL-719, Inavolisib/GDC-0077 and AZD-5363 treatment

CC cell lines, including CaSki (ATCC Cat# CRM-CRL-1550_Ca Ski, RRID: CVCL_1100), SiHa (ATCC Cat# HTB-35_SiHa, RRID: CVCL_0032 and ME180 (ATCC Cat# HTB-33_ME-180, RRID: CVCL_1401), C33A (ATCC CRM-HTB-31, RRID:CVCL_1094) were obtained from ATCC. SNU-17 (RRID: CVCL_5029) and SNU-902 (RRID: CVCL_5107) were kindly provided by the Korean Cell Line Bank ^40^. Other cell lines, including CERV-186 (CVCV_5729), were obtained from Cytion, and JHUCS-3 (CVCL_4653) from the RIKEN Bioresource Center. Cells were cultured according to the provider’s instructions and were regularly tested for mycoplasma infection and their identity by Identifiler. BYL-719, a potent and selective PI3Kα (*PIK3CA/*p110) inhibitor with IC of 5nM, was purchased from MCE (Cat# HY-15244, MedChem Express). GDC-0077 and AZD-5363 were also purchased from MedChem Express, All drugs were dissolved in DMSO, stored at -80EC, and diluted with an appropriate concentration before use according to the manufacturer’s protocol.

### Cervical cancer data analysis

TCGA (The Cancer Genome Atlas) data sets, including 297 cervical cancers (CESC), AACR-GENIE (AACR Project Genomics Evidence Neoplasia Information Exchange), including 190 CC, were accessed through the cBioPortal for Cancer Genomics analytic tool [40] ; A dataset of 22 CC cell lines. The 297 TCGA-CESC samples included 251 SCC, 46 ACC, and 178 samples with gene-mutation and HPV-type data, respectively. Of 194, the *PIK3CA* gene has the highest rate of mutations, and of 178, 103 HPV16 includes 78 with integration and 25 unintegrated tumors.

### Survival analysis

#### TCGA cohort

Only those with squamous cell carcinoma were examined to determine the effect of *YAP1* amplification on survival. The resulting dataset contained 29 individuals with *YAP1* amplification and 197 without amplification. A Kaplan-Meier survival analysis was conducted to observe the crude effect of *YAP1* amplification on survival. Cox proportional hazards ratios were calculated to assess the impact of YAP1 amplification on survival, adjusting for stage, tumor grade, age, and race.

Caris cervical Cancer patients were selected based on the absence of pathogenic or likely pathogenic mutation in *PIK3CA* or *STK11*. Overall survival was examined in the patient sub-cohort which was stratified based on the presence (green) of absence (yellow) of YAP1 amplification. Overall survival (OS) was defined as the time from the index date (sample collection) to death or last clinical contact. For patients with a recorded date of death, OS was calculated as the number of days from sample collection to death. For patients without a recorded death date, OS was calculated as the number of days from sample collection to the last clinical contact, defined as the maximum date observed across all claims sources and the collection date. Patients without a recorded death date were classified as having an inferred death if their last clinical contact occurred more than 100 days prior to the global last contact date in the dataset. If there was no recorded or inferred death, then patients were censored at the date of last clinical contact. Overall survival was estimated using the Kaplan-Meier method and compared across defined cohorts of the analysis population. Survival and treatment curves are visualized with 95% confidence intervals and risk tables generated to display patient numbers and events over time. Differences in survival between defined cohorts were examined using Cox proportional hazards regression to estimate hazard ratios (HRs). Multivariate analysis was performed to adjust for age and genomic ancestry with the sub population.

### Western Blot Analysis

According to the manufacturer’s protocols, whole-cell protein lysates were prepared from cell lines using RIPA lysis buffer supplemented with Complete Protease Inhibitor Cocktail (Santa Cruz, CA, United States). In total, 20 - 40 micrograms of whole cell protein were separated in a 4–12% NuPAGE Bis-Tris-gel, transferred to PVDF membrane (Thermo Fisher Scientific, Carlsbad, CA, United States), and immunoblotted with primary antibodies in 0.05% Tween 20-Tris-buffered saline (TBST) containing 5% skim milk at 4°C with shaking overnight. The primary antibodies used in this study were: rabbit monoclonal anti-PD-L1, (Cell Signaling Technology Cat# 13684, RRID: AB_2687655), YAP1, (Cell Signaling Technology Cat# 14074, RRID: AB_2650491), EGFR (Cell Signaling Technology Cat# 4267, RRID: AB_2800085), CTGF (Cell Signaling Technology Cat# 86641, RRID: AB_2800085), Integrin α6 (Cell Signaling Technology Cat# 3750, RRID: AB_2249263), IRF1 (Cell Signaling Technology Cat# 8478, RRID: AB_10949108) at a dilution of 1:500; mouse monoclonal anti-HPV16 E7 at 1:100 (Santa Cruz Cat# sc-51951, RRID: AB_629662), and goat monoclonal anti-p16 at 1:500 (R&D system Cat# AF5779, RRID: AB_1964666). β-actin, used as an internal control, was detected by rabbit anti-β-actin antibody (Cell Signaling Technology Cat# 4967, RRID: AB_330288). Anti-rabbit IgG, HRP-linked Antibody (Cell Signaling Technology Cat# 7074, RRID: AB_2099233) and Anti-mouse IgG, HRP-linked Antibody at 1:3000 - 1:5000 (Cell Signaling Technology Cat# 7076, RRID: AB_330924) were used as a secondary antibody. The membranes were incubated with SuperSignal West Duration Substrate (Thermo Scientific, United States) for 2 min, and the membranes were analyzed using the ChemiDoc Image System (Bio-Rad). Densitometric analysis of specific signals using Image Lab software 6, on three independent blots, was quantified and normalized to loading control β-actin.

The digital images were generated by confirming the correct molecular weight of the targeted proteins. For instance, HPV16 E7, a protein with a small molecular weight (less than 20 kDa), was analyzed. The blots were cut at the 30 kDa marker or higher, based on the protein marker (Novex™ Sharp Pre-stained Protein Standard, Cat. LC5800, Invitrogen) used. Digital images for the same cell line were typically produced from 1–2 membranes. Additional western blots were performed if more target proteins required analysis, ensuring consistent conditions throughout.

### Long-read RNA sequencing

Total RNA from CaSki cells treated with or without BYL-719 was extracted using an RNA Mini Kit (QIAGEN) according to the manufacturer’s protocol. The quality and quantity of RNA were assessed using a NanoDrop spectrophotometer, an Agilent 2100 Bioanalyzer, and the Qubit™ RNA Broad Range (B.R.) assay on a Qubit® Fluorometer (Invitrogen). Then, 50 ng of total RNA was subjected to RNA sequencing using the PCR-cDNA Barcoding kit (SQK-CB109; Oxford Nanopore Technologies (ONT)). Sequencing was performed on a MinIon Flow Cell using a GridIon DNA Sequencer (ONT), with base calling performed on the instrument. After obtaining cleaned reads, each sample was aligned to the exome and the compiled reference genome with Minimap2, using the Exome module of EPI2ME (ONT) [41].

### Quantitative TaqMan real-time PCR

The TaqMan RT-PCR method was used to confirm the result of gene expression from RNA sequence, Briefly 900ng total RNA was synthesized cDNA using random hexamer primer from TaqMan Reverse Transcription Reagents (Thermo Fisher) according to the manufacturer’s instruction and real-time (RT) PCR performed using TaqMan® Gene Expression Assays consist of a 20X mix of primers and TaqMan® MGB probe (FAM™ dye-labeled) in a single-plex PCR Reaction Mix using TaqMan® Universal PCR Master Mix in QuantStudio^TM^ Real-Time PCR system (software v1.7.2) (Thermo Fisher) [42]. The primer and probes are listed in **Supplemental Table 4.**

### Protein expression and cell proliferation in CC cell lines treated with BYL-719

CaSki, ME180, SNU-17, and SiHa were cultured in 6-well plates with 3.5×10^5^/well and treated with BYL-719 at 5µM, 10µM, 25µM, 50µM, then the cells were harvested after culturing for 24h and 48h. Protein levels were detected using Western blot as described in the Western Blot analysis section above. According to the provider’s protocol, the Agilent xCELLigence RTCA DP Instrument was used to measure cell proliferation plots. The xCELLigence RTCA was placed in a humidified incubator at 37 °C and 5% CO2 for real-time monitoring of cell proliferation^43^. Briefly, 100 µl of culture medium was added to each well of the 16-well E-plates (E-plate16) to measure background. Subsequently, 100 µl of cells (2 × 10^5/mL) were seeded in quadruplicate into each well for impedance-based detection. Sterile water was added to the outside chamber to maintain humidity. Further CaSki cells were seeded at 2×10^4^ cells per well and treated with BYL-719 0µM as a control and 5µM at first 24h; the following 24h and 48h the cultured medium was replaced with medium only (control), 0µM (5µM to 0µM), 1µM (5µM to 1µM), and 5µM (5µM to 5µM), respectively. Meanwhile, the same cells were cultured in 6-well plates for phase-contrast imaging at 72h and harvested at 72h for Western blotting. The cell index was monitored every 30 minutes for at least 72 hours, and data were recorded using the supplied RTCA software.

### Combination Cytolytic Assay: NEXI-003 and BYL-719

T cells specific against HR HPV tumor antigens from HLA A*02:01 healthy donor PBMCs were enriched and expanded using the Nexlmmune Artificial Immune Modulation (AIM) nanoparticle platform [32]. Briefly, AIM nanoparticles function as artificial antigen-presenting cells (APCs) that present antigenic peptides in the context of HLA class I and simultaneously deliver a second co-stimulatory signal, thereby engaging and expanding antigen-specific T cells for adoptive cell therapy (ACT). Different batches of antigen-specific T cells (research scale NEXI-003 products) derived from healthy donors were tested. The day prior to testing, T cells were thawed and recovered overnight from cryopreservation in TexMACS medium (Miltenyi Biotec) with cytokine supplements [43]. CaSki and HeLa-A2 (both HPV HLA A*02) were treated with either BYL-719 (5 µM) or T cells alone, or with both concomitantly. In BYL-719 wash out experiment, CaSki cells were pretreated with 5 µM BYL-719 for 24h followed by medium refreshing without or with BYL-719 (1 µM). Effector T cells (E:T ratio - 4:1) were also added at the time of medium refreshing.

xCELLigence RTCA was used to measure tumor cell proliferation. Cell counts (Mean ± SEM) are relative to 0h, just before the additions of either BYL-719 or T cells. The concentration and time that cells were treated with either BYL-719 or T cells alone or both in combination at the effector cell-to-target cell ratios (E:T ratios) of 2.5:1-10:1 are indicated in the figures.

### Statistics and Calculations

Mann-Whitney-U and two-tailed t-test and Pearson’s correlation analysis were performed using GraphPad Prism version 9 (RRID: SCR_002798) for Windows; p<0.05 was considered statistically significant. A one-way ANOVA was performed using GraphPad Prism version 9 (P < 0.05). Survival analyses were performed using R version 4.4.0. Percent inhibition was calculated from quadruplicate measurements of cell index as measured by the Agilent xCELLigence RTCA DP Instrument. For each condition, the values used were the maximum cell growth values from the control group to provide the most accurate estimate of inhibition relative to untreated control cells. Percent inhibition values were analyzed using an ordinary one-way ANOVA test assuming a Gaussian distribution (α = 0.05). Homogeneity of variances was verified by Brown-Forsythe and Bartlett’s tests. Since each treatment group was compared to a single control, post hoc multiple comparisons were performed using Dunnett’s test. All statistical analyses and percent inhibition figure creation were performed using GraphPad Prism Version 10.6.1.

For IC calculations, cell viability was quantified using real-time cell index measurements and expressed as percent viability relative to vehicle-treated controls. For each experiment, cell index values were extracted at a single fixed time point corresponding to the same elapsed time post-treatment across all concentrations for a given cell line. This timepoint was selected to align with the interval used for percent inhibition analyses and occurred prior to growth plateauing.

Viability values were normalized such that the mean response at 0 µM corresponded to 100%. Dose-response relationships were modeled using nonlinear regression in R with the drc package. IC values were estimated by fitting a two-parameter log–logistic model with fixed upper (100%) and lower (0%) asymptotes. Only non-zero drug concentrations were included in model fitting. Effective doses corresponding to 50% inhibition (ED) and associated 95% confidence intervals were calculated using the delta method. Experiments exhibiting non-monotonic dose–response behavior were excluded from IC estimation.

## Data Availability

All data produced in the present work are contained in the manuscript

## Acknowlegements

We thank Janina Pearce, Qian Wang, and Katherine Stang for comments on the manuscript. This research was supported [in part] by the Intramural Research Program of the National Institutes of Health (NIH). The contributions of the NIH author(s) are considered Works of the United States Government. The findings and conclusions presented in this paper are those of the author(s) and do not necessarily reflect the views of the NIH or the U.S. Department of Health and Human Services.

## Author Contributions

Conceptualization, H.L., D.L., M.O. and M.D.; methodology, H.L., D.L., N.S., E.A.M., S.T., T.J., I.R., S.K., E.R., N.M.R, Y.X., and N.R. writing—original draft preparation, H.L.; writing—review and editing, D.L., E.R., N.S, S.T., M.R.O., M.O. and M.D. All authors have read and agreed to the published version of the manuscript.

## Availability of data and material

Not applicable

